# Literature analysis of the efficacy of COVID-19 vaccinations

**DOI:** 10.1101/2022.12.24.22283835

**Authors:** Tim Hulsen

## Abstract

The COVID-19 pandemic is the largest epidemic of the 21^st^ century so far. Over 650 million people have already been infected with the SARS-CoV-2 virus. One of the ways to stop this pandemic, is to vaccinate the population and gain herd immunity. Many different vaccines are being used around the world, with differing efficacy. This review summarizes the 79 publications on the efficacy of the currently existing COVID-19 vaccines. It shows that there are eleven vaccines that have efficacy data published in a PubMed-indexed scientific journal. Most research has been done on the Pfizer/BioNTech BNT162B2 vaccine, and the eleven vaccines generally have a high efficacy in preventing illness. The Pfizer (86%-100%), Moderna (93.2%-94.1%), Sputnik-V (91.6%) and Novavax (∼90%) vaccines show the highest efficacy, followed by the Sinovac (83.5%), QazCovid-in 82%) and Covaxin (77.8%) vaccines. The Oxford/AstraZeneca (69% - 81.5%) and Johnson & Johnson (66%) vaccines have lower efficacy in preventing illness. This overview also shows efficacies other than in preventing illness (e.g. asymptomatic, severe illness, hospitalization, death) in some cases. The results also show that the vaccines have specific effects on specific age groups (e.g. adolescents, adults, elderly) and people with diseases (e.g. leukemia, other cancers, HIV). Future research in this area will mostly focus on vaccine efficacy on specific strains of the SARS-CoV-2 virus (such as the Omicron variant) as well as the efficacy of booster vaccinations.

## 1. Introduction

The COVID-19 pandemic is the largest epidemic of the 21^st^ century so far. As of December 19, 2022, 658 million people have contracted the Severe Acute Respiratory Syndrome CoronaVirus 2 (SARS-CoV-2 virus)[1]. There are several ways to prevent the spread of COVID-19, the disease caused by SARS-CoV-2 infection: e.g. by washing hands, wearing face masks, practicing social distancing, staying home when sick (quarantine) and by vaccinations. The latter option does not only work on short term, but also on the longer term, by creating ‘herd immunity’ if the larger part of a population has become immune to the virus. Development of vaccines was started immediately after the sequencing of the SARS-CoV-2 virus in January 2020 [2]. In September 2020, 10 SARS-CoV-2 vaccine candidates were already in phase III clinical trials [3]. At this moment, many different vaccines are being used around the world, with differing efficacy. Much research on COVID-19, SARS-CoV-2 vaccines and their efficacy has been done already [4]. A review of the four most used COVID-19 vaccines in August 2021 (‘BNT162b2/Pfizer’, ‘mRNA-1273/Moderna’, ‘Oxford/AstraZeneca/AZD1222’ and ‘Janssen/Ad26.COV2.S’), including their efficacy and geographical distributions, was published by Francis et al. [5]. They reported high efficacy for the Pfizer (95%) and Moderna (94.1%) vaccines and lower efficacy for the AstraZeneca (70.4%) and Janssen (66.9%) vaccines. Another review by Fiolet al. [6] showed that the Pfizer, Moderna and Sputnik V vaccines had the highest efficacy (>90%) after two doses. Rotschild et al. [7] also offered a comparison of some of the mostly widely used vaccines and concluded that the mRNA vaccines (Pfizer and Moderna) were associated with the highest efficacy in preventing symptomatic COVID-19. The data used in these reviews are all obtained from efficacy studies done previously. However, at this moment more vaccines are available, and more vaccine efficacy studies have been published in the meantime. This review summarizes the 79 publications available in PubMed on the efficacy of the currently existing COVID-19 vaccines, providing us with an overview of the current status of available vaccines and how effective they are. It also shows information on the efficacy in specific age groups and patient groups with diseases. Finally, it will give some directions for future research around SARS-CoV-2 vaccine efficacy.

## 2. Materials and Methods

### 2.1. Search strategy

A list of available vaccines was retrieved from Our World In Data [8,9]. PubMed was then queried for each vaccine, using the search term “(sars-cov-2 OR covid-19[ti]) vaccine[ti] efficacy[ti]”, in combination with the vaccine-specific search term with the producer, product name and scientific name(s), such as “(bnt162b2[ti] OR pfizer[ti] OR biontech[ti] OR comirnaty[ti])”, on 2022-12-19 (figure 1).

**Figure 1.**
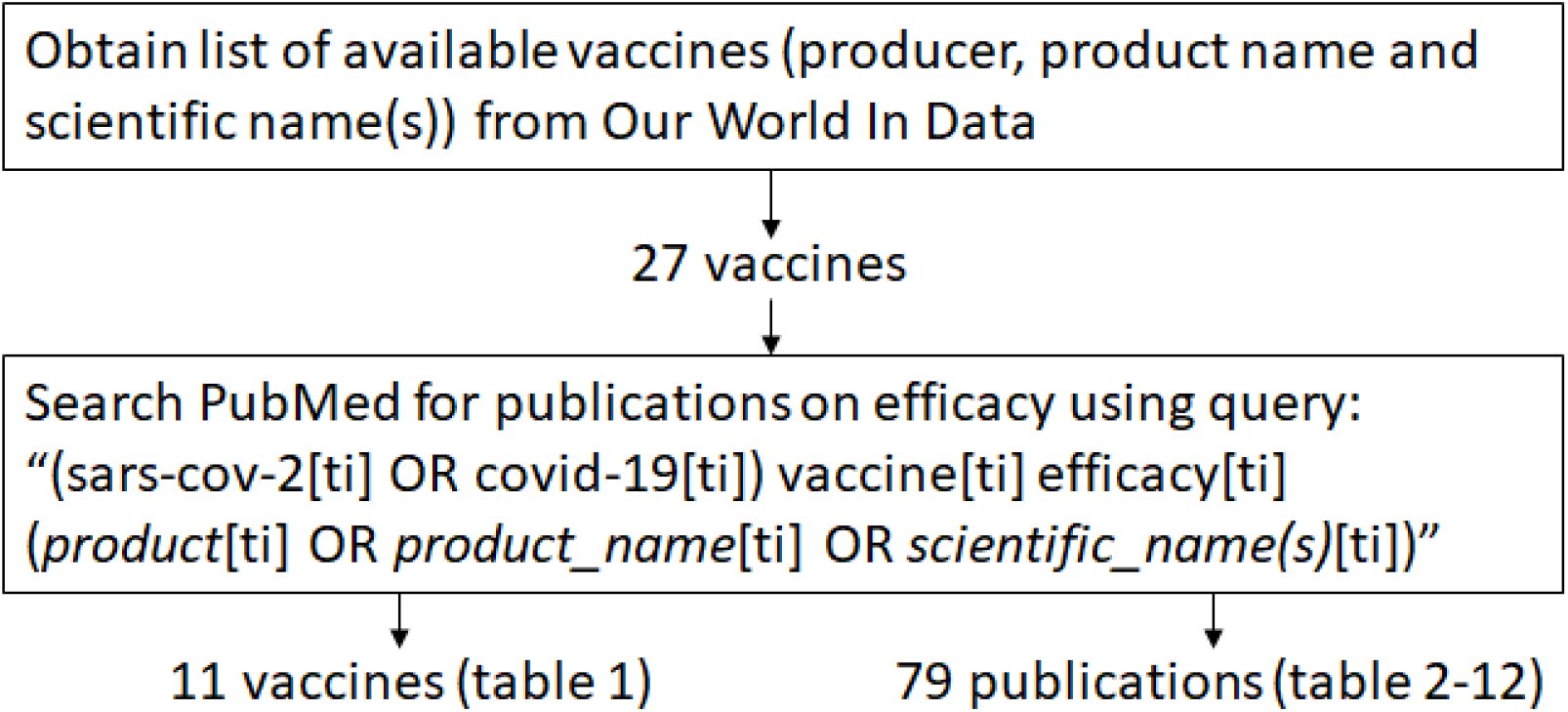
Flow chart describing the search strategy.

### 2.2 Data processing

Results were collected in an Excel sheet, with the literature data for each vaccine on a separate tab. These data were then used to create tables 1-12. Abstracts as well as full-text manuscripts were analyzed to summarize the findings, displayed in the Results section. Because of the large number of publications for the Pfizer/BioNTech vaccine, the text was split up into several subsections (general efficacy, efficacy in adolescents, efficacy in CLL patients, efficacy in other cancer patients, efficacy in patients receiving haemodialysis, efficacy in other patients). Vaccines that did not have any publications on efficacy were not included in the tables. One publication [10] contained information on two vaccines.

**Table 1.** Overview of COVID-19 vaccines.

| Producer | Country of origin | Product name | Scientific name(s) | Type | # Pubs |
| --- | --- | --- | --- | --- | --- |
| Pfizer/BioNTech | USA/Germany | Comirnaty | BNT162B2 | mRNA | 32 |
| Oxford/AstraZeneca | UK/Sweden | Covishield/Vaxzevria | ChAdOx1 | vector | 11 |
| Moderna | USA | Spikevax | mRNA-1273, mRNA1273 | mRNA | 9 |
| Novavax | USA |  | NVX-CoV2373 | protein subunit | 8 |
| Johnson & Johnson | Netherlands/Belgium/USA | Janssen | Ad26.COV2.S | vector | 7 |
| Covaxin | India |  | BBV152 | inactivated | 3 |
| Sinovac | China | CoronaVac |  | inactivated | 3 |
| Sinopharm/Beijing | China | Sinopharm BIBP | BBIBP-CorV | inactivated | 3 |
| Gamaleya | Russia | Sputnik V | Gam-COVID-Vac, rAd26, rAd5 | vector | 2 |
| CanSino | China | Convidecia | AD5-nCOV | vector | 1 |
| QazCovid-in | Kazakhstan | QazVac |  | inactivated | 1 |

Vaccines obtained from Our World In Data [8], with the number of publications on efficacy in PubMed.

## 3. Results

### 3.1. Available vaccines

The full list of available vaccines, with publications on efficacy in PubMed (as per 2022-12-19), is shown in table 1. It also shows the country of origin, the product name(s) and scientific name. The type can be “mRNA”, “vector”, “protein subunit”, “inactivated” or “peptide subunit”. The rightmost column shows the number of publications that were found using the PubMed query mentioned in section 2.1.

### 3.2. The Pfizer/BioNTech BNT162B2 vaccine

The Pfizer/BioNTech vaccine has the most publications of all (32, table 2). There are 14 publications about the general efficacy, 2 about efficacy of the vaccine in adolescents, and 16 about efficacy in specific patient groups: 3 about chronic lymphocytic leukemia (CLL) patients, 7 about other cancer patients, 2 about patients receiving haemodialysis, and 4 in other patients.

**Table 2.**
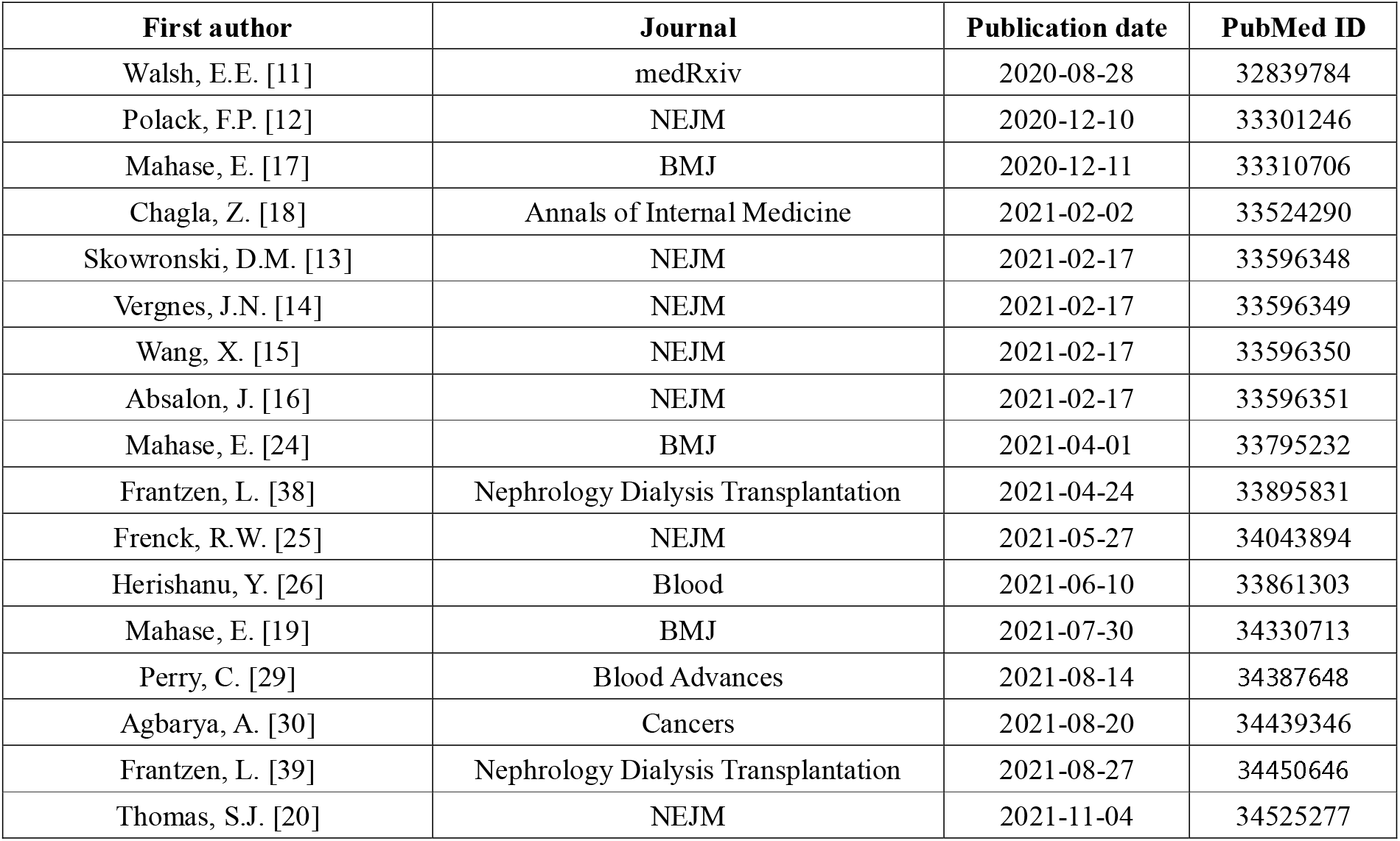
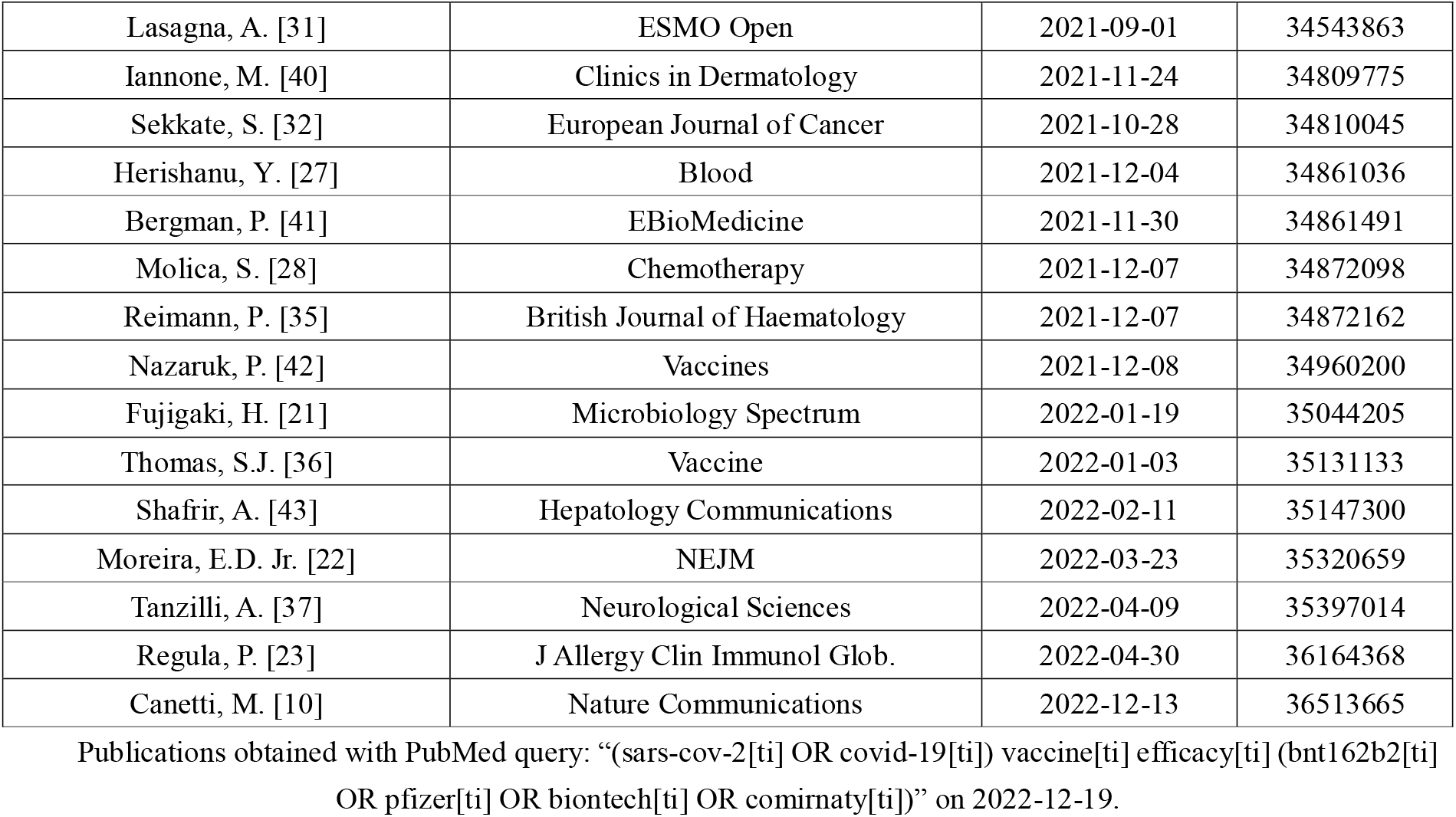
Publications on the efficacy of the Pfizer/BioNTech BNT162B2 vaccine.

#### 3.2.1. General efficacy of the Pfizer/BioNTech BNT162B2 vaccine

The first publication was a preprint of Walsh et al. [11], which researched both the Pfizer/BioNTech BNT162b1 and BNT162b2 vaccine candidates. Their results showed that, in both younger (18–55 years old) and older (65–85 years old) adults, the two vaccine candidates caused similar dose-dependent SARS-CoV-2-neutralizing geometric mean titers (GMTs), comparable to or higher than the GMT of a panel of SARS-CoV-2 convalescent sera. The BNT162b2 vaccine candidate was associated with less systemic reactogenicity, particularly in 65-85 years olds.

The second publication about efficacy (as well as safety) of the Pfizer/BioNTech BNT162b2 mRNA COVID-19 vaccine was Polack et al. [12]. They concluded that a 2-dose regimen of BNT162b2 gave 95% protection against COVID-19 in persons >= 16 years old. This study was then followed-up in 2021 with three comments and the authors’ reply. Comment #1 from Skowronski & De Serres [13] said that due to vaccine shortage in the USA, second doses should be postponed. Comment #2 from Vergnes [14] pointed to discrepancies in the table with age groups, and that this should have been clarified. Comment #3 from Wang [15] stated that the preliminary data do not support the conclusion that this vaccine offers protection against severe COVID-19 illness, since the percentage of COVID-19-positive patients with severe illness was significantly higher in the vaccine group compared to the placebo group. The authors replied [16] that 1) alternative dosing regimens of BNT162b2 had not been evaluated, and that the decision to implement alternative dosing regimens resides with health authorities and not the authors (working at Pfizer); 2) Vergnes incorrectly summed the COVID-19 cases in the age groups, and the assertion that the data overestimate vaccine efficacy in the age groups is unsubstantiated; and 3) it is not appropriate to use the proportion of COVID-19-positive patients in whom severe disease developed to assess vaccine protection against severe COVID-19. The results of the Polack et al. publication [12] were referenced in two articles published by Mahase [17] and Chagla [18], focusing on the results that the Pfizer vaccine efficacy was 52% after first dose and 95% after second dose. However, Mahase later on reported that the efficacy of the BNT162b2 vaccine dropped from 96% to 84% four months after second dose [19]. Thomas et al. [20] reported an efficacy of 91.3% after 6 months of follow-up. There was a gradual decline in vaccine efficacy. Vaccine efficacy of 86% to 100% was seen across populations. Vaccine efficacy against severe disease was 96.7%. In South Africa, where the SARS-CoV-2 variant B.1.351 (‘Beta’) was predominant, a vaccine efficacy of 100% was observed.

Fujigaki et al. [21] analyzed the levels of the immunoglobulins IgG, IgM, and IgA against the receptor-binding domain (RBD) of SARS-CoV-2 in the sera from 219 people in Japan who received two doses of the BNT162b2 vaccine. They showed that there were much higher levels of RBD-IgG after the second vaccination. The IgG levels significantly correlated with the neutralizing activity. They concluded that the monitoring of IgG against RBD is a powerful tool to predict the vaccination efficacy and provides useful information in considering a personalized COVID-19 vaccination strategy.

Moreira et al. [22] assessed vaccine safety and efficacy against COVID-19 starting 7 days after the third (booster) dose in persons 16 years of age or older, in a clinical trial in which 5081 participants received a third BNT162b2 vaccine and 5044 received a placebo. They conclude that a third dose of the BNT162b2 vaccine administered ∼11 months after the second dose provided 95.3% efficacy.

Regula et al. [23] researched the efficacy of the Pfizer-BioNTech vaccine on subjects experiencing immediate allergic reactions to the first dose. Their findings indicate, by measuring COVID-19 spike antibody levels, that graded dosing of this vaccine is efficacious and useful for treating these subjects with allergy.

Canetti et al. [10] report a three-month follow-up of 700 participants in a fourth vaccine dose study, comparing the Pfizer/BioNTech BNT162b2 and Moderna mRNA-1273 vaccines, administered four months after a third BNT162b2 dose. Both vaccines show little efficacy against infection but were highly efficacious against substantial symptomatic disease (71% for BNT162b2 and 89% for mRNA-1273).

#### 3.2.2. Efficacy of the Pfizer/BioNTech BNT162B2 vaccine in adolescents

Two articles reported results from studies in specific age groups. Mahase [24] published results from a Pfizer press release reporting that as shown 100% efficacy against SARS-CoV-2 in 12 to 15 year olds in the preliminary results of a phase III trial. Frenck et al. [25] indeed confirmed these results from the clinical trial with 2260 adolescents. Apparently, the efficacy of the Pfizer/BioNTech BNT162B2 vaccine is especially high in adolescents.

#### 3.2.3. Efficacy of the Pfizer/BioNTech BNT162B2 vaccine in CLL patients

Sixteen more articles focused on the efficacy of the BNT162b2 vaccine in patients with specific diseases. Herishanu et al. [26] studied the efficacy of the vaccine on patients with CLL, which have an increased risk for severe COVID-19 disease and mortality. Their results show that antibody-mediated response to the BNT162b2 mRNA COVID-19 vaccine in patients with CLL is markedly impaired and affected by disease activity and treatment. A follow-up study by the same authors [27] showed that patients with CLL/small lymphocytic lymphoma (SLL) who failed to achieve a humoral response after standard two-dose BNT162b2 mRNA vaccinations, close to 25% responded to the third dose of vaccine. Molica et al. [28] assessed humoral immune and cellular responses to the BNT162b2 messenger RNA (mRNA) COVID-19 vaccination in CLL. They concluded that serological response to the BNT162b2 vaccine in patients with CLL is impaired, and that a third vaccine dosage should be considered for these patients.

#### 3.2.4. Efficacy of the Pfizer/BioNTech BNT162B2 vaccine in other cancer patients

Perry et al. [29] published their findings on the efficacy of the BNT162b2 vaccine in patients with B-cell non-Hodgkin lymphoma (B-NHL). They concluded that patients treated with an anti-CD20 antibody were unlikely to achieve humoral response to the vaccine, and that longer time since last exposure to anti-CD20 antibodies predicts a higher response rate and elevated antibody titer.

Agbarya et al. [30] analyzed the humoral response following vaccination with the second dose of BNT162b2 in patients with solid malignancies who were receiving anti-cancer therapy at the time of vaccination. The humoral response in the cancer patient group was significantly lower than in the non-cancer group. They concluded that reduced immunogenicity to the vaccine among chemotherapy-treated cancer patients, raises the need to continue exercising protective measures after vaccination in these patients.

Lasagna et al. [31] aimed to assess both humoral and cellular response after a messenger RNA (mRNA) vaccination schedule. They demonstrated that the administration of a full course of BNT162b2 vaccine elicited a sustained immune response against SARS-CoV-2 regardless of the type of cancer and/or immune checkpoint inhibitors.

Sekkate et al. [32] commented on Shmueli et al. [33], a paper describing that adequate antibody response after BNT162b2 vaccination was achieved after two doses but not after one dose, in 129 patients with cancer vaccinated during anticancer therapy. Sekkate et al. state that Shmueli et al. investigated the clinical characteristics potentially associated with seronegativity, including age, BMI, type of cancer, stage, comorbid conditions and type of treatment, but did not take the degree of lymphopenia into account. According to their results, there is a significant correlation between antibody and lymphocyte levels. Therefore, lymphocyte counts appear to be a simple potential criterion that could be used to improve the selection of patients requiring an additional booster dose in the future. Shacham-Shmueli et al. [34] responded with a post-hoc analysis, which suggested that low lymphocyte counts following the second BNT162b2 vaccine dose correlate with low antibody titers. They concluded that the presence of low lymphocyte count may be taken into consideration, among other factors, in decision-making for administering a third (booster) vaccine dose.

Reimann et al. [35] investigated efficacy and safety of a heterologous booster vaccination with Ad26.COV2.S DNA vector vaccine in haemato-oncological patients without antibody response after double-dose BNT162b2 vaccine. Ad26.COV2.S is the Johnson & Johnson vaccine, and will be discussed in that section.

Thomas et al. [36] report results from 3,813 participants with a history of past or active neoplasm and up to 6 months’ follow-up post-dose 2 from the placebo-controlled, observer-blinded trial of the 2-dose BNT162b2 mRNA COVID-19 vaccine. Vaccine efficacy was 94.4%, compared to a vaccine efficacy of 91.1% in the overall trial with 46,429 participants.

Tanzilli et al. [37] studied the safety and efficacy of the Pfizer/BioNTech vaccine in patients affected by primary brain tumor (PBT). They examined 112 patients at the Neuro-oncology Unit of the Regina Elena National Cancer Institute, of which 102 received a first dose, 100 received the second one and 73 patients received the booster dose. After the first dose they observed one patient with fever and severe fatigue. After the second dose they recorded adverse events in ten patients. No correlation was observed between adverse events and comorbidities.

#### 3.2.5. Efficacy of the Pfizer/BioNTech BNT162B2 vaccine in patients receiving haemodialysis

Frantzen et al. [38,39] presented their results on a group of patients receiving haemodialysis, who have a particular high mortality rate (close to 20%) when infected by SARS-CoV-2. They reported that their patients, after receiving two injections (3 weeks apart) of the BNT162b2 vaccine, had no serious adverse events. 91% presented a positive antibody titer. Older patients were less likely to present an antibody response. There was also no response to vaccination among all patients undergoing chemotherapy or under immunosuppression.

#### 3.2.6. Efficacy of the Pfizer/BioNTech BNT162B2 vaccine in other patients

Iannone et al. [40] presented a case report on the safety and efficacy of the BNT162b2 vaccine during Ixekizumab treatment for hidradenitis suppurativa (HS) on a 48-year-old woman. Their case confirmed that vaccine administration in a patient with HS treated with anti–IL-17 drugs is effective. The patient developed high-titered antibody response, despite the important role of Th17 axis in natural infection clearance and in the development of vaccine-induced immunity.

Bergman et al. [41] investigated safety and efficacy of BNT162b2 mRNA vaccination in five selected groups of immunocompromised patients (primary immunodeficiency disorders, secondary immunodeficiency disorders due to human immunodeficiency virus infection, allogeneic hematopoietic stem cell transplantation/CAR T cell therapy, solid organ transplantation (SOT), or chronic lymphocytic leukemia (CLL)). and healthy controls. Their results showed that the efficacy of the mRNA BNT162b2 vaccine was lower in immunocompromised patients compared to healthy controls. They highlight the need for additional vaccine doses in certain immunocompromised patient groups to improve immunity.

Nazaruk et al. [42] determined the immune response to the BNT162b2 vaccine in 65 kidney transplant recipients (KTRs) and 65 liver transplant recipients (LTRs). Patients in both cohorts received two 30 μg doses of BNT162b2 vaccine in 3-to-6-week intervals. Their results showed a higher than previously reported humoral response to the BNT162b2 vaccine in KTRs and LTRs, which was dependent upon age, type of transplanted organ, and immunosuppression.

Shafrir et al. [43] assessed the impact of hepatic fibrosis on the efficacy of the Pfizer/BioNTech vaccine. They concluded that immune suppression, older age, male gender, and advanced chronic liver disease are risk factors for lower vaccine response, and that the Fibrosis-4 (FIB-4) score can be used to prioritize candidates for third-dose vaccine booster.

### 3.3. The Oxford/AstraZeneca ChAdOx1 vaccine

Eleven articles were published on the Oxford/AstraZeneca ChAdOx1 vaccine (table 3). Mahase [44] presents an early (June 2020) report on the development of the Oxford/AstraZeneca vaccine, stating that trials were started in Brazil and South Africa, with another country in Africa set to follow, as well as a trial in the US. The manuscript explains that these trials can be accelerated because existing technology is used, and researchers are using only one of the genes to make one of the proteins from the pathogen. Next to this vector vaccine, the author also briefly discusses ongoing work on mRNA vaccine candidates as well as vaccines created with an inactivated version of SARS-CoV-2 and a live attenuated version.

**Table 3.**
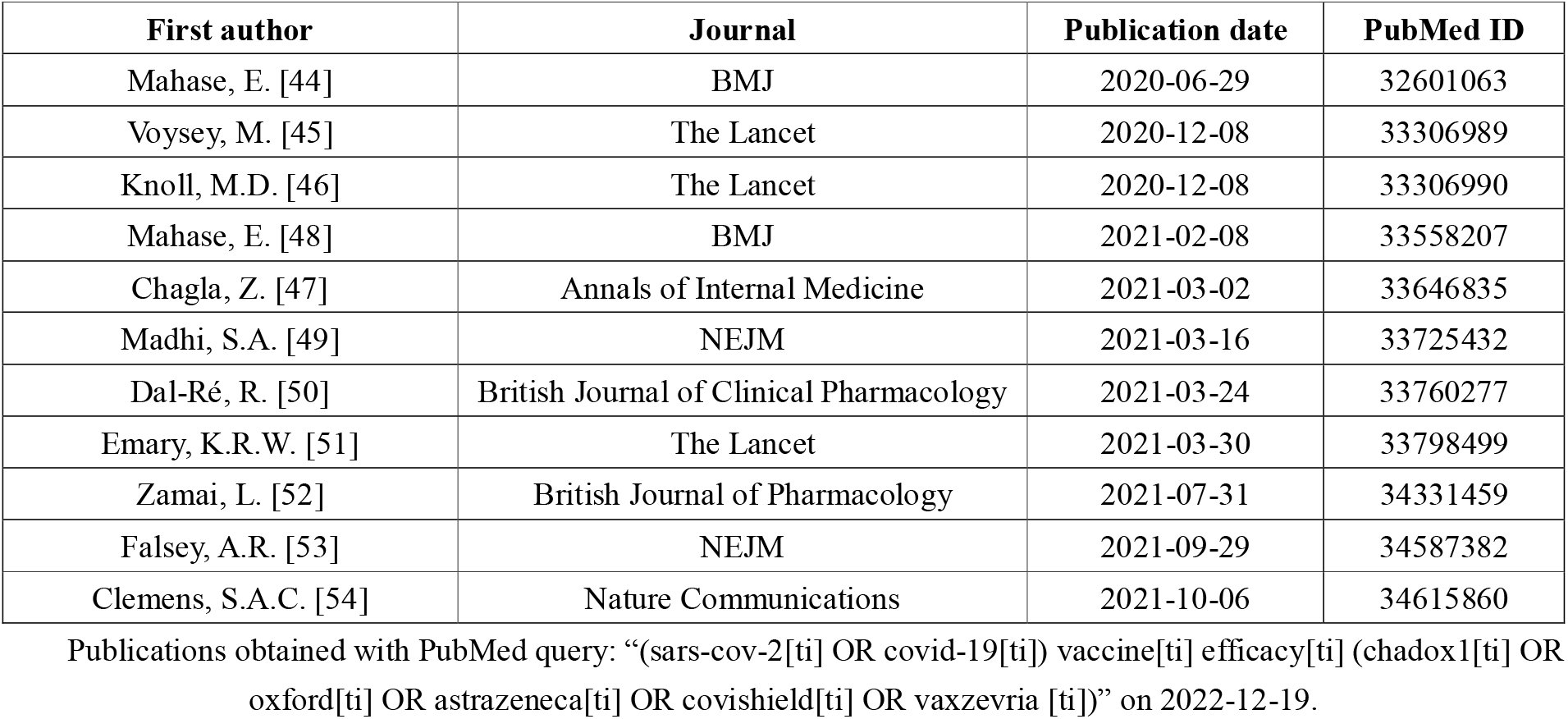
Publications on the efficacy of the Oxford/AstraZeneca ChAdOx1 vaccine.

In December 2020, the results of the randomised controlled trials mentioned above (in Brazil, South Africa, and the UK) were presented by Voysey et al. [45]. 23,848 participants were enrolled and 11,636 participants were included in the interim primary efficacy analysis. In participants who received two standard doses, vaccine efficacy was 62.1% and in participants who received a low dose and a standard dose, efficacy was 90.0%. Overall vaccine efficacy across both groups was 70.4%. From 21 days after the first dose, there were 10 cases from the control arm hospitalised for COVID-19, with one leading to death. This paper was commented on by Knoll and Wonodi [46], who welcome the publication because it is the first report of efficacy for a non-profit COVID-19 vaccine aiming for global supply, equity, and commitment to poorer countries. They point to the strengths of the study, such as the large sample size, diversity of the patients, balance of participant characteristics between the vaccine groups, and having similar results in Brazil as in the UK, lending credibility to the results. According to Knoll and Wonodi, the study also has some limitations: less than 4% of participants were over 70 years old, no participants older than 55 years received the mixed-dose regimen, and those with comorbidities were a minority, with results for that subgroup not yet available. Chagla [47] summarizes the trial results and compares them with those of the Moderna and Pfizer/BioNTech vaccine trials. He points to the advantage of the Oxford/AstraZeneca vaccine that it can be stored at 2 to 8 °C. According to him, a major concern in the approval of the Oxford/AstraZeneca vaccine is how to process the inconsistencies in dosing, since 2 dosing regimens were used in the trials.

Another publication by Mahase [48] in February 2021 brings the news that the rollout of the Oxford-AstraZeneca COVID-19 vaccine in South Africa was paused after a study in 2026 healthy and young volunteers reported that it did not protect against mild and moderate disease caused by the B.1.351 (501Y.V2, ‘Beta’) variant which was already accounting for 90% of cases in South Africa at the time. Efficacy against severe COVID-19, hospital admissions, and deaths were not determined. Other vaccines showed lower efficacy against the Beta variant as well. These results were published by Madhi et al. [49] one month later. They showed that mild-to-moderate COVID-19 developed in 3.2% of the placebo-receiving cohort and in 2.5% of the vaccine-receiving cohort, leading to an efficacy of 21.9%. Among the 42 participants with COVID-19, 39 cases were caused by the B.1.351 variant; vaccine efficacy against this variant was 10.4%.

Dal-Ré [50] stated in March 2021 that the first efficacy data from clinical trials on the Oxford/AstraZeneca vaccine did not include many older adults (≥56 years old), which is why, at the time, the vaccine was not approved yet in some countries, such as Switzerland and the USA. Data from an ongoing trial would show the efficacy of the vaccine in older adults, as well as data from countries that did approve the vaccine already.

Emary et al. [51] shows the efficacy of the Oxford/AstraZeneca vaccine against the B.1.1.7 (‘Alpha’) variant. 8534 participants were included in the trial, of which 520 participants developed SARS-CoV-2 infection. Clinical vaccine efficacy against symptomatic nucleic acid amplification test (NAAT) positive infection was 70.4% for B.1.1.7 and 81.5% for non-B.1.1.7 lineages. They conclude that the Oxford/AstraZeneca vaccine showed reduced neutralisation activity against the B.1.1.7 variant compared with a non-B.1.1.7 variant in vitro, but the vaccine showed efficacy against the B.1.1.7 variant of SARS-CoV-2.

Zamai et al. [52] suggests that mass vaccination with the Oxford/AstraZeneca vaccine would not quickly solve the pandemic, as anti-vector immunity takes place as homologous vaccination with the Oxford/AstraZeneca vaccine is applied and interferes with vaccine efficacy when the interval between prime and booster doses is less than three months. Overmore, this vaccine might provide suboptimal efficacy against the B.1.1.7 (‘Alpha’) variant (as discussed above), which appears to have an increased transmissibility among vaccinated people. They suggest physical preventive measures as well as the adoption of an influenza-like vaccination strategy.

In September 2021, Falsey et al. [53] presented the results of a phase 3 clinical trial on 32,451 participants (including many older adults, lacking in previous trials) on the Oxford/AstraZeneca vaccine. Vaccine efficacy was 83.5% in participants 65 years of age or older, and the overall estimated vaccine efficacy was 74.0%. High vaccine efficacy was consistent across a range of demographic subgroups. In the fully vaccinated analysis subgroup, no severe or critical symptomatic COVID-19 cases were observed among the 17,662 participants, while 8 cases were noted among the 8,550 participants in the placebo group (<0.1%). The estimated vaccine efficacy for preventing SARS-CoV-2 infection was 64.3%.

One month later, Clemens et al. [54] investigated the efficacy of the Oxford/AstraZeneca vaccine against symptomatic COVID-19 in a post-hoc exploratory analysis of a Phase 3 randomised trial in Brazil. Protection against any symptomatic COVID-19 caused by the P.2 (‘Zeta’) variant was assessed in 153 cases with vaccine efficacy of 69%. 49 cases of B.1.1.28 (a predominant variant in Brazil) occurred and vaccine efficacy was 73%. The P.1 (‘Gamma’) variant arose later in the trial and only 18 cases were available for analysis; vaccine efficacy was 64%. The Oxford/AstraZeneca vaccine provided 95% protection against hospitalisation due to COVID-19.

### 3.4. The Moderna mRNA-1273 vaccine

Nine articles were published on the Moderna mRNA-1273 vaccine (table 4). Mahase [55] summarizes a press release from Moderna, stating that results were released from its phase III trial (the COVE study), showing vaccine efficacy of 94.1% based on 196 covid-19 cases, of which 185 were in the placebo group. These results were then published by Baden et al. [56] and summarized a few months later by Chagla et al. [57]. El Sahly et al. [58] present results from further analyses of efficacy and safety data from the blinded phase of the trial. Vaccine efficacy in preventing COVID-19 illness was 93.2, the efficacy in preventing severe disease was 98.2%, and the efficacy in preventing asymptomatic infection starting 14 days after the second injection was 63.0%. Vaccine efficacy was consistent across ethnicities, age groups, and participants with coexisting conditions. Follmann et al. [59] finally evaluated antinucleocapsid antibody (anti-N Ab) seropositivity in mRNA-1273 vaccinees with breakthrough SARS-CoV-2 infection, in a substudy of the COVE study. Tré-Hardy et al. [60] performed a study in which they compared the antibody responses at three time points (T1: 2 weeks after the first injection, T2: 2 weeks after the second injection, T3: 3 months after the first injection) from 205 healthcare workers stratified according to their initial serological status. They evaluated the immune response of the participants but also the effectiveness of the Moderna mRNA-1273 vaccine in a context different from the previous studies by Moderna. At T3, they confirmed a very high efficacy and a persistence of anti-spike antibodies.

**Table 4.** Publications on the efficacy of the Moderna mRNA-1273 vaccine.

| First author | Journal | Publication date | PubMed ID |
| --- | --- | --- | --- |
| Mahase, E. [55] | BMJ | 2020-12-02 | 33268462 |
| Baden, L.R. [56] | NEJM | 2020-12-30 | 33378609 |
| Chagla, Z. [57] | Annals of Internal Medicine | 2021-03-02 | 33646836 |
| Tré-Hardy, M. [60] | Journal of Infection | 2021-06-20 | 34161817 |
| Gilbert, P.B. [62] | medRxiv | 2021-08-15 | 34401888 |
| El Sahly, H.M. [58] | NEJM | 2021-09-22 | 34551225 |
| Gilbert, P.B. [61] | Science | 2021-11-23 | 34812653 |
| Follmann, D. [59] | Annals of Internal Medicine | 2022-07-05 | 35785530 |
| Canetti, M. [10] | Nature Communications | 2022-12-13 | 36513665 |

Publications obtained with PubMed query: “(sars-cov-2[ti] OR covid-19[ti]) vaccine[ti] efficacy[ti] (moderna[ti] OR spikevax[ti] OR mrna-1273[ti] OR mrna1273[ti])” on 2022-12-19.

Gilbert et al. [61] (preprint: [62]) present the results of the COVE phase 3 clinical trial, in which vaccine recipients were assessed for neutralizing and binding antibodies as correlates of risk for COVID-19 disease and as correlates of protection. Vaccine recipients with postvaccination 50% neutralization titers 10, 100, and 1000 had estimated vaccine efficacies of 78%, 91%, and 96%, respectively.

Canetti et al. [28] was discussed in the section Pfizer/BioNTech BNT162b2 section above.

### 3.5. The Novavax NVX-CoV2373 vaccine

Eight articles were published on the Novavax NVX-CoV2373 vaccine (table 5). Mahase [63] discusses preliminary results from clinical trials, showing that the Novavax vaccine is 95.6% effective against the original variant of SARS-CoV-2 but also provides protection against the B.1.1.7 variant (‘Alpha’, 85.6%) and B.1.351 variant (‘Beta’; 60%). These data were obtained from three ongoing clinical trials: one in the UK, one in the USA and Mexico (both phase III) and one in South-Africa (phase II).

**Table 5.** Publications on the efficacy of the Novavax NVX-CoV2373 vaccine.

| First author | Journal | Publication date | PubMed ID |
| --- | --- | --- | --- |
| Mahase, E. [63] | BMJ | 2021-02-01 | 33526412 |
| Shinde, V. [64] | NEJM | 2021-05-05 | 33951374 |
| Heath, P.T. [65] | NEJM | 2021-06-30 | 34192426 |
| Sacks, H.S. [66] | Annals of Internal Medicine | 2021-11-02 | 34724399 |
| Toback, S. [67] | The Lancet Respiratory Medicine | 2021-11-17 | 34800364 |
| Granwehr, B.P. [68] | Annals of Internal Medicine | 2022-05-03 | 35500261 |
| Montastruc J.L. [70] | Fundamental & Clinical Pharmacology | 2022-05-02 | 35502459 |
| Heath, P.T. [71] | Clinical Infectious Diseases | 2022-10-10 | 36210481 |

Publications obtained with PubMed query: “(sars-cov-2[ti] OR covid-19[ti]) vaccine[ti] efficacy[ti] (novavax[ti] OR nvx-cov2373[ti])” on 2022-12-19.

Shinde et al. [64] discusses the results of a South African phase 2a-b trial, with HIV-positive and HIV-negative participants. They concluded that the Novavax vaccine was efficacious in preventing COVID-19, with higher vaccine efficacy (60.1%) observed among HIV-negative participants. The B.1.351 variant, which likely originated in South-Africa, caused most of the infections.

Heath et al. [65] presents a phase 3, randomized, observer-blinded, placebo-controlled trial with 14,039 participants from 33 sites in the UK. Infections were reported in 10 participants in the vaccine group and in 96 in the placebo group, with a symptom onset of at least 7 days after the second injection, for a vaccine efficacy of 89.7%. No hospitalizations or deaths were reported among the 10 cases in the vaccine group. Five cases of severe infection were reported, all in the placebo group. The Novavax vaccine had an efficacy of 86.3% against the B.1.1.7 variant and 96.4% against other variants. Sacks [66] comments on these results, stating that their data corresponds well with preliminary unpublished data from a larger Novavax trial, still ongoing in the USA and Mexico, which also found 90% efficacy against symptomatic COVID-19. He notes that, as of August 2021, there is little information on the Novavax vaccine’s effectiveness against the B.1.617.2 (‘Delta’) variant. Overmore, the Novavax vaccine might be particularly attractive to developing countries, because it only requires refrigeration (not freezer storage), just like the Johnson and Johnson Ad26.COV2.S vaccine.

There is limited data on efficacy of COVID-19 vaccines combined with seasonal influenza vaccines. Toback et al. [67] therefore reported results of a substudy within a phase 3 UK trial, by evaluating the safety, immunogenicity, and efficacy of NVX-CoV2373 when co-administered with licensed seasonal influenza vaccines. They concluded that co-administration resulted in no change to influenza vaccine immune response although a reduction in antibody responses to the NVX-CoV2373 vaccine was noted. NVX-CoV2373 vaccine efficacy in the substudy was 87.5% and in the main study was 89.8%.

Granwehr [68] discusses another paper by Dunkle et al. (2022) [69] on the efficacy and safety of the NVX-CoV2373 vaccine in adults in the United States and Mexico. The authors concludes that the vaccine efficacy was 67% in Hispanic/Latino participants compared with 90% overall, which confirms the findings of other recent studies that there is considerable variation in efficacy according to race or ethnicity. Since many clinical trials show low diversity of race or ethnicity, this makes it difficult to characterize potentially important differences in vaccine response.

Montastruc et al. [70] performed some more analysis on the data from Heath et al. [65]. They calculated three different values for risk expression: absolute risks (AR), absolute risk reduction (ARR), number needed to treat (NNT) and relative risks (RR). ARR and NNT values were 1.22% and 82, respectively, for an RR value of 0.10. AR values were 0.14% in exposed patients and 1.36% in placebo patients. They state that the low AR in placebo patients could explain the difficulties of perception of COVID-19 risks by some parts of the population. They also conclude that, taking into account ARR and NNT, the NVX-Cov2373 vaccine seems to be more active in younger adults than in older adults, and the RR values indicate that the NVX-Cov2373 vaccine failed to show any clinical efficacy in non-white patients, in contrast to Caucasians.

Heath et al. [71] present the final results of the aforementioned phase 3 trial in the UK. Of the 13,989 remaining subjects, 6,989 received the NVX-CoV2373 vaccine and 7,000 the placebo. At a maximum of 7.5 months postvaccination, there were 24 cases of COVID-19 among NVX-CoV2373 recipients and 134 cases among placebo recipients, a vaccine efficacy of 82.7%. Vaccine efficacy was 100% against severe disease and 76.3% against asymptomatic disease.

### 3.6. The Johnson & Johnson Ad26.COV2.S vaccine

Seven articles were published on the efficacy of the Johnson & Johnson Ad26.COV2.S vaccine (table 6). Sadoff et al. [72] reported the results of a clinical trial with 39,321 participants. They found out that a single dose of the Ad26.COV2.S vaccine protected against symptomatic COVID-19 and asymptomatic SARS-CoV-2 infection and was effective against severe-critical disease, including hospitalization and death. Vaccine efficacy with onset at least 14 days after administration was 66.9% and for onset at least 28 days after administration it was 66.1%. It was higher against severe-critical COVID-19 (76.7% for ≥14 days and 85.4% for ≥28 days). Heininger [73] commented on this publication, suggesting that their statement that “the data do not suggest a waning of protection” is misleading, as without sufficient exposure to severe acute respiratory syndrome coronavirus (SARS-CoV-2), waning of immunity cannot be detected clinically. He states that more data are needed to reliably assess the duration of protection after a single dose of the Ad26.COV2.S vaccine. The authors replied [74] that they only intended to contextualize that vaccine efficacy over time was driven mainly by smaller groups of participants who were followed for long periods of time, in whom no evidence of waning efficacy was observed up to day 91. They agree that additional and longer-term data are needed to assess the durability of protection.

**Table 6.**
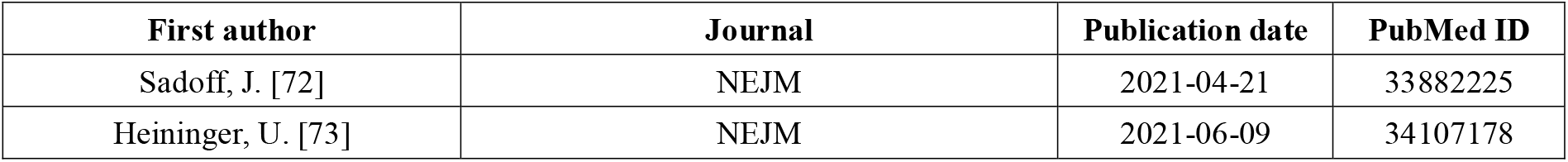
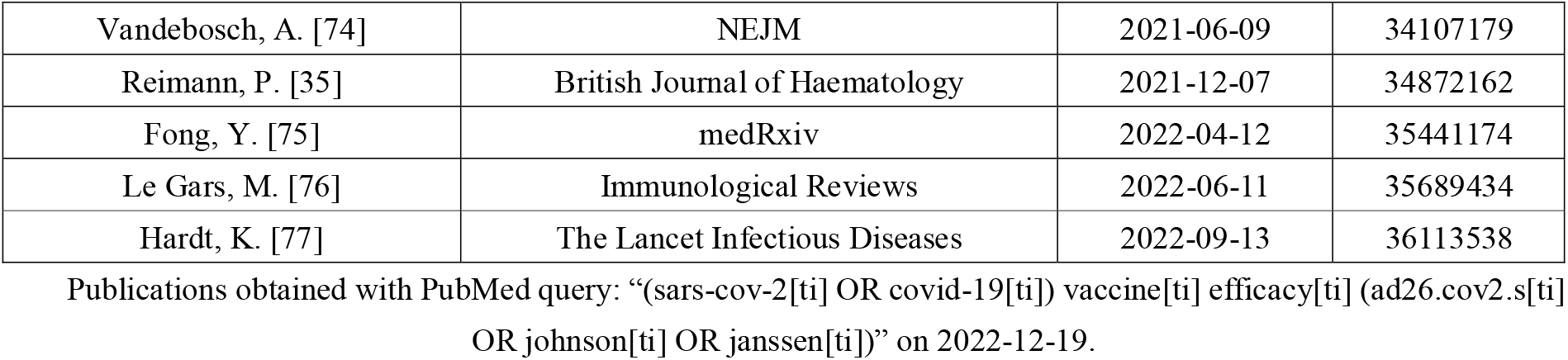
Publications on the efficacy of the Johnson & Johnson Ad26.COV2.S vaccine.

Reimann et al. [35] investigated efficacy and safety of a heterologous booster vaccination with Ad26.COV2.S DNA vector vaccine in haemato-oncological patients without antibody response after double-dose BNT162b2 vaccine. This vaccination led to a serological response in 31.0% of the patients without response after double-dose BNT162b2. They concluded that this vaccination regimen should be further evaluated to increase the response rate in the highly vulnerable population of haemato-oncological patients.

Fong et al. [75] published a preprint with the results from the ENSEMBLE trial. In this international, randomized efficacy trial, antibody measurements 28 days after vaccination were assessed as correlates of risk of moderate to severe-critical COVID-19 outcomes through 83 days after vaccination and as correlates of protection following a single dose of the Johnson & Johnson Ad26.COV2.S vaccine. Each marker had evidence as a correlate of risk and of protection, with strongest evidence for 50% inhibitory dilution (ID50) neutralizing antibody titer. Vaccine efficacy was 60% at nonquantifiable ID50 (< 2.7 IU50/ml) and rose to 89% at ID50 = 96.3 IU50/ml.

Le Gars et al. [76] discusses the efficacy and immunogenicity of Ad26.COV2.S as a single-dose primary vaccination and as a (homologous or heterologous) booster vaccination. They found that this vaccine causes broad humoral and cellular immune responses, which are associated with protective efficacy/effectiveness against SARS-CoV-2 infection, moderate to severe/critical COVID-19, and COVID-19-related hospitalization and death, including against emerging SARS-CoV-2 variants (e.g. Delta and Omicron).

Hardt et al. [77] present the results of the ENSEMBLE2 trail, a randomised, double-blind, placebo-controlled, phase 3 trial including crossover vaccination after emergency authorisation of COVID-19 vaccines, with 31,300 participants from 10 countries. Vaccine efficacy was 75.2% against moderate to severe-critical COVID-19. Most cases were due to the variants alpha (B.1.1.7) and mu (B.1.621).

### 3.7. The Covaxin BBV152 vaccine

Three articles were published on the efficacy of the Covaxin BBV152 vaccine (table 7), of which two in animal models and one in humans. Mohandas et al. [78] studied the immunogenicity and efficacy of inactivated SARS-CoV-2 vaccine candidates BBV152A, BBV152B, and BBV152C in Syrian hamsters (*Mesocricetus auratus*). BBV152A and BBV152B vaccine candidates remarkably generated a quick and robust immune response. The protective efficacy of all three candidates was demonstrated by lower viral load, absence of lung pathology, and high titers of neutralizing antibodies post-infection, with BBV152A showing the best response.

**Table 7.** Publications on the efficacy of the Covaxin BBV152 vaccine.

| First author | Journal | Publication date | PubMed ID |
| --- | --- | --- | --- |
| Mohandas, S. [78] | iScience | 2021-01-09 | 33521604 |
| Yadav, P.D. [79] | Nature Communications | 2021-03-02 | 33654090 |
| Ella, R. [80] | The Lancet | 2021-11-11 | 34774196 |

Publications obtained with PubMed query: “(sars-cov-2[ti] OR covid-19[ti]) vaccine[ti] efficacy[ti] (covaxin[ti] OR bbv152[ti])” on 2022-12-19.

Yadav et al. [79] developed and measured the protective efficacy and immunogenicity of the same three candidate vaccines in rhesus macaques (*Macaca mulatta*). Twenty macaques were divided into four groups of five animals each. One group was administered a placebo, while three groups were immunized with the vaccine candidates at 0 and 14 days. All animals were infected with SARS-CoV-2 14 days after the second dose. The protective response was observed with increasing SARS-CoV-2 specific IgG and neutralizing antibody titers from 3rd-week post-immunization. Viral clearance was observed at 7 days post-infection in the three vaccinated groups.

Ella et al. [80] present the interim results of the phase 3 trial of BBV152 on 25,798 participants. The overall estimated vaccine efficacy was 77.8%. The vaccine was well tolerated since the same proportion of participants reported adverse events in the vaccine group and placebo group.

### 3.8. The Sinovac CoronaVac vaccine

Three articles were published on the efficacy of the Sinovac CoronaVac vaccine (table 8). Palacios et al. [81] describes an ongoing phase 3 clinical trial in Brazil to assess the efficacy and safety of the adsorbed vaccine COVID-19 (inactivated) produced by Sinovac. Two age groups are investigated: adults (18-59 years old) and elderly (≥60 years old). The participants are receiving two intramuscular doses of either the CoronaVac vaccine or a placebo (randomized, 1:1 ratio). The primary efficacy endpoint is the incidence of symptomatic cases of virologically confirmed COVID-19 two weeks after the second vaccination. The virological diagnosis will be confirmed by detection of SARS-CoV-2 nucleic acid in a clinical sample.

**Table 8.** Publications on the efficacy of the Sinovac CoronaVac vaccine.

| First author | Journal | Publication date | PubMed ID |
| --- | --- | --- | --- |
| Palacios, R. [81] | Trials | 2020-10-15 | 33059771 |
| Tanriover, M.D. [82] | The Lancet | 2021-07-08 | 34246358 |
| Jin, L. [83] | Human Vaccines & Immunotherapeutics | 2022-07-25 | 35878789 |

Publications obtained with PubMed query: “(sars-cov-2[ti] OR covid-19[ti]) vaccine[ti] efficacy[ti] (sinovac[ti] OR coronavac[ti])” on 2022-12-19.

Tanriover et al. [82] presents the results of a phase 3 clinical trial in Turkey in which 10,214 participants received two doses of the vaccine or a placebo (ratio approximately 2:1). During a median follow-up period of 43 days, nine cases of PCR-confirmed symptomatic COVID-19 were reported in the vaccine group and 32 cases were reported in the placebo group 14 days or more after the second dose, yielding a vaccine efficacy of 83.5%. They conclude that the CoronaVac vaccine has high efficacy against PCR-confirmed symptomatic COVID-19.

Jin et al. [83] summarized the results of clinical trials and real-world studies of the CoronaVac vaccine. The overall efficacy for the prevention of symptomatic COVID-19 (before the emergence of variants of concern) using two doses was 67.7%. Effectiveness in preventing hospitalizations, intensive care unit admissions, and deaths was more prominent than that in preventing COVID-19. A third dose inherited the effectiveness against non-variants of concern and increased effectiveness against severe COVID-19 outcomes caused by omicron variants compared to two doses.

### 3.9. The Sinopharm/Beijing BBIBP-CorV vaccine

Three publications show data on efficacy of the Sinopharm/Beijing BBIBP-CorV vaccine (table 9). Karimi et al. [84] studied the efficacy and side effects of the Sinopharm vaccine in 434 patients with hemoglobinopathies in Iran. They found that the efficacy was not optimal due to the lack of effect on new variations of the virus, but that the vaccine seemed to be protective against the severity of COVID-19 infection in hemoglobinopathy patients.

**Table 9.** Publications on the efficacy of the Sinopharm/Beijing BBIBP-CorV vaccine.

| First author | Journal | Publication date | PubMed ID |
| --- | --- | --- | --- |
| Karimi, M. [84] | Mediterr J Hematol Infect Dis. | 2022-03-01 | 35444764 |
| Ghasemi, A. [85] | Current Drug Safety | 2022-08-01 | 35927906 |
| Alirezaei, A. [86] | Iranian Journal of Kidney Disease | 2022-07-01 | 35962641 |

Publications obtained with PubMed query: “(sars-cov-2[ti] OR covid-19[ti]) vaccine[ti] efficacy[ti] (sinopharm[ti] OR beijing[ti] OR bbibp-covv[ti])” on 2022-12-19.

Ghasemi et al. [85] investigated the efficacy of the Sinopharm vaccine in 53 bone marrow transplant (BMT) recipients in Iran. They reported that vaccine side effects were reported in only 7.7% of the patients, and that vaccine failure against the Delta and Omicron variants was reported in 39.6% of the cases.

Alirezaei et al. [86] compared the level of anti-SARS-CoV-2 anti-spike protein receptor-binding domain IgG neutralizing antibody before and after vaccination with two doses of Sinopharm vaccine, in patients undergoing hemodialysis. The absolute mean change in antibody titer following full-scheduled vaccination was 8.98 μg/mL, and the rate of seroconversion was 31.1. The rate of seroconversion was higher in subjects who had COVID-19 previously than in those who did not have it.

### 3.10. The Sputnik-V Gam-COVID-Vac vaccine

Two articles show the efficacy of the Sputnik-V Gam-COVID-Vac vaccine (table 10). Logunov et al. [87] report preliminary results on the safety and efficacy of Gam-COVID-Vac from an interim analysis of a phase 3 trial on 21,977 participants. The vaccine efficacy was 91.6% after two doses. Tsimafeyeu et al. [88] describes the preliminary safety and efficacy of the Gam-COVID-Vac vaccine in patients with active genitourinary (GU) malignancies. They showed that patients with GU malignancies are at increased risk of mortality from COVID-19 infection when compared to the general population: the 3-month survival rate was only 82%.

**Table 10.** Publications on the efficacy of the Sputnik-V Gam-COVID-Vac vaccine.

| First author | Journal | Publication date | PubMed ID |
| --- | --- | --- | --- |
| Logunov, D.Y. [87] | The Lancet | 2021-02-20 | 33545094 |
| Tsimafeyeu, I. [88] | Journal of Hematology & Oncology | 2021-11-13 | 34774086 |

Publications obtained with PubMed query: “(sars-cov-2[ti] OR covid-19[ti]) vaccine[ti] efficacy[ti] (gamaleya[ti] OR sputnik-v[ti] OR gam-covid-vac[ti] OR rad26[ti] OR rad5[ti])” on 2022-12-19.

### 3.11. The CanSino AD5-nCOV vaccine

Only one article discusses the efficacy of the CanSino AD5-nCOV vaccine (table 11). Guzman-López et al. [89] carried out a retrospective cohort study on 7,468 teachers in Mexico, during the COVID-19 delta wave. Primary breakthrough symptomatic infections were higher in the CanSino vaccinated group compared to other brands. For CanSino vaccine recipients, anti-S antibodies were >50 AU/mL in 73.2% of the participants. No difference was found between CanSino and other vaccines regarding hospitalization, the need for mechanical ventilation, and death. Participants with a previous infection had higher antibody levels than those who were reinfected and without infection.

**Table 11.**
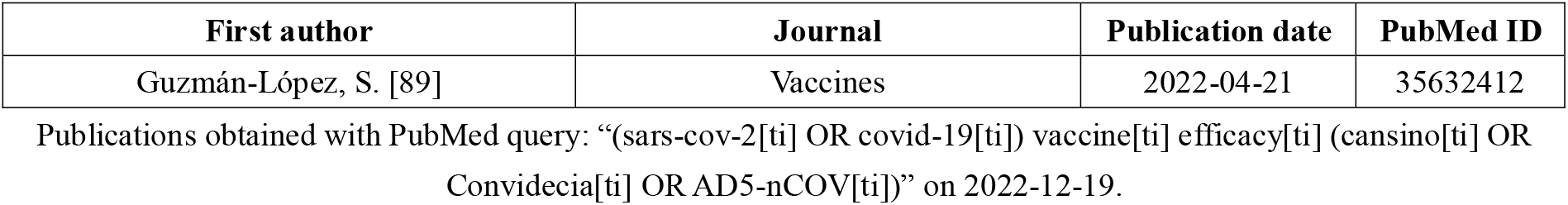
Publications on the efficacy of the CanSino AD5-nCOV vaccine.

### 3.12. The QazCovid-in QazVac vaccine

One manuscript presents results on the efficacy of the QazCovid-in QazVac vaccine (table 12). Khairullin et al. [90] conducted a multicenter, randomised, single-blind, placebo-controlled phase 3 trial of the QazCovid-in vaccine with a 180-day follow-up period in three clinical centres in Kazakhstan. Out of 2,400 vaccinated participants, 31 were diagnosed with COVID-19; 43 COVID-19 cases were recorded in 600 placebo participants with onset of 14 days after the first dose within the 180-day observation period. The protective efficacy of the QazCovid-in vaccine was 82.0%.

**Table 12.** Publications on the efficacy of the QazCovid-in QazVac vaccine.

| First author | Journal | Publication date | PubMed ID |
| --- | --- | --- | --- |
| Khairullin, B. [90] | eClinicalMedicine | 2022-06-25 | 35770251 |

Publications obtained with PubMed query: “(sars-cov-2[ti] OR covid-19[ti]) vaccine[ti] efficacy[ti] (qazcovid-in[ti] OR QazVac[ti])” on 2022-12-19.

## 4. Conclusions

The 79 publications summarized in this manuscript provide an insightful overview of the current status of the efficacy of the eight most prominent vaccines available. Several types of efficacies were studied (asymptomatic, preventing illness/severe illness/hospitalization/death), but usually the central research question is about the efficacy in preventing illness. By far the most data is available on the Pfizer/BioNTech vaccine, and these data show that this vaccine has a high efficacy (86% to 100%) in preventing illness, not only in the general population but also in adolescents, specific patient groups (patients with CLL or other cancers, patients receiving haemodialysis, and other patients). The Oxford/AstraZeneca vaccine has a lower efficacy, in the range of 69% - 81.5% and even (much) lower for the ‘Beta’ strain of SARS-CoV-2. The Moderna vaccine has an efficacy of 93.2% - 94.1% in preventing illness. The Novavax vaccine has an efficacy of around 90% for the original SARS-CoV-2 strain but lower for the newer ones. The Johnson & Johnson vaccine has an efficacy of around 66%. The Covaxin, Sinovac, Sputnik-V and QazCovid-in vaccines have efficacies of 77.8%, 83.5%, 91.6% and 82.0% respectively, based on single publications. Generally speaking, most vaccines have a high efficacy, especially for the original SARS-CoV-2 strain. The results also show that the vaccines have specific effects on specific age groups (e.g. adolescents, adults, elderly) and people with diseases (e.g. leukemia, other cancers, HIV). It should be noted that efficacy data at this moment is still limited and based on a small number of clinical trials. Some publications already showed some efficacy numbers for specific variants of the SARS-CoV-2 virus [20,49,51,65], but in general this subject needs much more future research. Data on the existing strains is still quite limited, and since the virus is still mutating, more strains might show up and cause resurgences of the COVID-19 pandemic. Therefore, it will be crucial that we will know what vaccines are effective against specific current and future SARS-CoV-2 strains, such as the Omicron variant (and its subvariants such as BA.4, BA.5 and BA.2.75 (‘Centaurus’) [91]). Some efficacy data for the Omicron variant are available for the Johnson & Johnson Ad26.COV2.S [76], Sinovac CoronaVac [83] and Sinopharm [85] vaccines, but the numbers of subjects are still relatively small, making it difficult to draw any conclusions. Moreover, some studies also measure the efficacy of multiple vaccinations and boosters [22,35]. Since the pandemic is still ongoing (especially in countries such as China), the efficacy of boosters will be important for the future as well.

## Data Availability

All data produced in the present work are contained in the manuscript.

## Acknowledgments

-

